# Familial Genetic Cancer Risk Assessment with respect to a Silent *BRCA2* mutation

**DOI:** 10.1101/2021.07.01.21258680

**Authors:** Ashwini Bapat, Siddharth Gahlaut, Rupa Mishra, Aijaz Ul Noor, Laleh Busheri, Ruhi Reddy, Shahin Shaikh, Ashraf Mannan, Smeeta Nare, Santosh Dixit, Chaitanyanand B. Koppiker

## Abstract

Male Breast Cancer (BC) is relatively rarer, accounting for less than 1% of cancers in men. MBC is hereditary in nature and mainly attributed to *BRCA1/2* germline mutations. Accordingly, National Comprehensive Cancer Network (NCCN) guidelines advise genetic counselling and testing for all cases of MBCs and their unaffected family members. In this report, we present an uncommon case of male patient primarily diagnosed with pancreatic cancer who later developed asynchronous bilateral hormone positive breast cancer. We describe the genetic screening and clinical management protocol for the proband and family members. Genetic testing with next generation sequencing by uses of a multi-gene germline mutation panel revealed a likely pathogenic *BRCA2* variant (c.8754G>A, p.E2918E). Subsequently, 34 members of the extended family of the proband were tested for the *BRCA2* variant by Sanger sequencing. 6 of the family members were identified as carriers of this *BRCA2* variant. Of these, three presented with hereditary breast cancer and 3 were unaffected healthy carriers. *In silico* analysis for mechanistic insights in underlying pathogenicity revealed that the silent *BRCA2* mutation is a spliceogenic variant that is likely to create an aberrant mRNA transcript via alternative splicing of *BRCA2* gene. Our study demonstrates the clinical relevance of this silent *BRCA2* mutation and emphasizes the need for further experimental studies to elucidate its functional role in breast cancer pathology.

## Introduction

Male Breast Cancer (MBC) is a rare disease accounting for 1% of the total breast cancers in the world (Moelans et al., 2019). The lifetime risk of a man developing breast cancer is 1:1000 (100 times less than that in a woman) (Abdelwahab Yousef, 2017). The risk of developing male breast cancer increases linearly after the age of 60 years and peaks around 70 years (Vietri et al., 2020) and is generally observed in older men. Given the rarity of the disease and lack of large cohorts, the recommendations and clinical management of the disease today, are mainly based on the data extrapolated from female breast cancer studies.

Demographic, environmental, epidemiologic and genetic factors are risk factors known to influence the onset of MBC (Weiss et al., 2005). Elevated levels of estrogen prior to radiation exposure and strong family history also contribute to the increased MBC risk (Piscuoglio et al., 2016). The NCCN guidelines for genetic testing of hereditary breast, ovarian, prostate and pancreatic cancers have suggested genetic testing in a breast cancer affected male as well as his 1^st^ degree relatives. MBCs are most frequently found to be Infiltrating Ductal Carcinoma and generally present as a lump in the breast. The staging of the cancer reflects the degree of tumour spread and the prognosis, in a fashion similar to breast cancer in females (Fentiman, 2016).

Pathogenic germline mutations in the high penetrance *BRCA1* and *BRCA2* genes, which act as tumour suppressors, have been long-established as the key mediators of predisposition to hereditary breast and ovarian cancers (Pfeffer et al., 2017). Functionally, *BRCA1* and *BRCA2* are key players in double stranded DNA repair pathways and maintain genomic integrity through homologous recombination during replication. Germline *BRCA1/2* mutations account for 25% of hereditary breast and ovarian cancer (HBOC). *BRCA2* contributes to about 6-10% of mutations in male breast cancer. Recent reports indicate that *BRCA1/2* mutations also increase the risk of developing pancreatic and prostate cancers (Ibrahim et al., 2018; Kast et al., 2016; Nielsen et al., 2016; Shah et al., 2020; Tripoli & Cordova, 2020).

The *BRCA1* gene with 24 exons, encodes an 1863 amino acid protein and is most frequently mutated in N-terminus RING domain, exon 11-13 and C-terminus BRCT domain (Pfeffer et al., 2017). The *BRCA2* with 27 exons, encodes for a 3418 amino acid protein and is characterised by the BRC repeat, a conservative segment, coded by exon 11, and a C-terminus oligonucleotide binding (OB) region (Venkitaraman, 2019).

Unlike the breast cancers in females, the patho-etiology of MBCs has not been well studied. Recent reports have highlighted an important role for *BRCA1/2* germline mutations in predisposition towards MBC. Population genetic studies have revealed that approximately 0-4% men diagnosed with breast cancer harbour a *BRCA1* mutation and 4-16% of them harbour a *BRCA2* mutation (Ferzoco & Ruddy, 2015). When compared with female breast cancers, *BRCA2* mutations are more common in MBCs than *BRCA1* mutations indicating the potential difference in etiology of breast cancers in males and females (Fentiman, 2016). MBCs with a *BRCA2* mutation present a significantly higher grade of breast cancers than *BRCA2* mutated cancers in females. The presentation of MBCs at an advanced stage is presumed to be due to lack of mammographic screening programs for men and the lack of awareness about MBCs (Silvestri et al., 2016).

In this report, we present a case of a male patient (proband) initially diagnosed with pancreatic cancer and subsequently diagnosed with asynchronous bilateral breast cancer. Genetic testing during clinical management identified the proband as a carrier of a *BRCA2* likely pathogenic variant (c.8754, G>A, p.E2918E). We present the clinical management and genetic screening approach for familial genetic risk assessment in the proband and his family. Furthermore, we provide insights into the potential biological mechanisms underlying the pathogenicity of this uncommon variant causing a synonymous change in *BRCA2* protein (p.E2918E).

## METHODOLOGY

### Data Collection

Detailed compilation of clinical meta-data of the male patient (proband) was undertaken from our clinic. As per NCCN guidelines, 34 consenting members of the extended family of proband, 2 of which were breast cancer affected, were included in familial genetic risk assessment protocol. Family pedigree and medical history was obtained from the family members. Raw data for sequencing was obtained from partner genetic testing laboratory while the clinicopathology features were obtained from the medical records. Written informed consent was obtained from all the individuals. The study was approved by the Institutional Ethics Committee of PCCM (DCGI/CDSCO Registration Number: ECR/298/Indt/MH/2018. Every participant gave a separate informed consent for the use of their personal and medical information associated with this study. The study follows principles of the declaration of Helsinki (Brazil, 2013 version) and National Ethics Guidelines 2017 issued by the Indian Council of Medical Research, Government of India.

### Clinical Management

The proband reported to our clinic with an initial diagnosis of pancreatic cancer and underwent clinical management in partner hospital. Breast Cancer diagnosis was based upon clinical examination and radiological evaluation of breast and axilla using CT abdomen 3 phase/ pelvis, Full Field Digital Mammography (FFDM) with 3-D Tomosynthesis (Siemens Mammomat Inspiration), Ultrasound, F18-FDG Whole Body PET CT scan and Ultrasonography (Siemens Acuson S2000). Histopathological studies on Tru-Cut biopsy on the sample was performed for confirming diagnosis of breast carcinoma. Similarly, ultrasonography and fine needle aspiration cytology were used for investigating axillary lymph node metastasis. The oncologic management with chemoradiation protocols was undertaken by a multidisciplinary clinical team in accordance with the current NCCN guidelines.

### Genetic Risk Assessment

The pre and post-test counselling was performed as per the NCCN guidelines. After informed consent, the genetic testing was performed as per American College of Medical Pathologists (ACMG) guidelines in CAP-certified partner genetic testing laboratories. For familial risk assessment (i.e., cascade testing), individual counselling sessions were conducted prior to and after the cascade testing reports were available.

### Genetic Testing

#### A. Next Generation Sequencing (NGS)

Genomic deoxyribonucleic acid (gDNA) was extracted from blood/saliva sample using standardized methodology (PrepIT-L2P kit from DNA Genotek, Canada) or QIAamp DNA Mini Kit (Qiagen, Germany) or the Nucleospin kit (Macherey–Nagel, Germany) and quantified by Qubit fluorimeter (Life Technologies). The next generation sequencing performed for the index patient included the genes associated with hereditary cancers and as per the ACMG guidelines. The TruSight Cancer Sequencing panel evaluated 30 genes associated with hereditary breast and endocrine cancers for the index patient. Next generation sequencing libraries were prepared by tagmentation, adaptor ligation, hybridization and were quantified to 6-10pM. Sequencing was performed using a standard v2 kit on Illumina MiSeq and the genetic variations were identified by using customised bioinformatics pipelines at partner genetic testing laboratory.

#### B. Sanger Sequencing

Sanger sequencing was performed in all 34 members of probands family for the variant c.8754G>A (p.E2918E), in exon 21 of the *BRCA2* gene. Genomic DNA extracted from blood/saliva was processed using big dye terminator sequencing kit (Lifetech Inc., USA). Capillary electrophoresis performed on the Genetic Analyzer (3500 DX, Lifetech Inc., USA) was visualized using the Finch TV software (Geospiza, USA). Presence of the variant, c.8754G>A (p.E2918E), was evaluated by comparing the sample sequence with the reference sequence (RefSeq id: NM_000059).

### *In Silico* Analysis

*In silico* splice site prediction tools namely NNSPLICE and Splice Port were utilized for bioinformatic analysis of spliceogenicity of the reported *BRCA2* variant and associated pathogenicity.

For identifying population prevalence and clinical significance of the *BRCA2* variant, database searches were undertaken for the reported *BRCA2* variant. These included online databases namely, CIMBA (Consortium of Investigators of Modifiers of *BRCA1/2*); ENIGMA (Evidence-based Network for the Interpretation of Germline Mutant Alleles); BCAC (The Breast Cancer Association Consortium); BIC (Breast Information Core); ClinVar and BRCA Exchange.

The family pedigree was constructed using Progeny (Progeny software LLC, South Bend, IN, USA).

## RESULTS

### Case Description

A male in his fifties presented with CT obstructive jaundice with peri-ampullary mass/ nodule measuring 1.4 x 2.2 cm. The mass was later confirmed to be pancreatic cancer by duodenoscopy-guided biopsy as a well differentiated adenocarcinoma of peri-ampullary region of pancreas. Whipple pancreaticoduodenectomy surgery was performed as part of clinical management.

A year later, the patient presented with a lump in one of his breast which was concluded to be benign epithelial lesions after a USG-guided FNAC procedure. However, the PET scan and subsequent biopsy revealed a Grade II Infiltrating Ductal Carcinoma. Immunohistochemical analysis identified the mass with an estrogen receptor (ER) / progesterone receptor (PR) positive (ER-80%, PR-70%), Human epidermal growth factor receptor (Her2-) negative status. Modified radical mastectomy was performed post primary diagnosis. The axillary excision biopsy revealed 2 out of 15 pathologically positive axillary lymph nodes, suggesting tumour metastases. Histopathology reports indicated moderately differentiated adenocarcinoma with perineural invasion and positive for Cytokeratin-7. Patient was treated with 6 cycles of 5-FluoroUracil; Epirubicin; Cyclophosphamide (FEC) adjuvant chemotherapy post-surgery. Subsequently, Linear accelerator based IMRT/3DCRT radiotherapy was administered. Hormonal therapy (Tamoxifen) was prescribed for 5 years after the completion of the treatment.

Two years later, the patient, presented with a bloody discharge from the nipple of the contralateral breast. On targeted sonomammography, two dilated ducts were seen at the 6 ‘o’clock position. The larger lesion was 2.7 mm in size. Internal echoes, increased peri ductal and internal vascularity was noted. Cytology of the nipple discharge of the was performed and was found to be positive for malignant cells. The biopsy findings and histological examinations characterised the breast tumour as Invasive ductal carcinoma Grade I, ER/PR positive. PET scan revealed a distant metastatic lesion in the left anterior abdominal wall measuring 61.5 x 29.9 mm. The patient was treated with Neoadjuvant hormone therapy (Anastrazole) and GnRH analogue which was followed by modified radical mastectomy.

In the same year, one of the relatives of the proband was diagnosed with ER/PR positive breast cancer (left). A few months later another relative was diagnosed with ER/PR positive breast cancer (right). In parallel, the proband was diagnosed with lung metastases and was treated with chemotherapy.

Considering the case history (i.e., pancreatic cancer followed by asynchronous bilateral breast cancer) was indicative of hereditary breast and ovarian cancer syndrome, the proband and family members were advised genetic testing. After written informed consent, the proband underwent a comprehensive multi-gene panel covering 30 genes associated with hereditary breast and ovarian cancer, including the 25 genes recommended by ACMG.

### Genetic Analysis by Multi-gene Panel

Multi-gene panel testing by next generation sequencing included 30 high, moderate and low penetrance genes recommended by ACMG, predisposing to HBOC, pancreatic cancer and prostate cancer. Interestingly, only one heterozygous ‘likely pathogenic’ variant (c.8754G>A) on exon 21 of a high penetrance *BRCA2* gene was reported out of the 30 genes tested.

The reported variant showed a heterozygous nucleotide change ‘G>A’ at position c.8754 in the *BRCA2* gene and was also confirmed by sequencing with forward primer (flanking the *BRCA2* variant) in two independent experiments. The detected variant resulted in no amino acid change (p.E2918E) when compared with the reference sequence (RefSeq ID: NM_000059). Since this variant was identified to cause a synonymous substitution in the *BRCA2* protein (p.E2918E), it was referred to as a silent mutation.

### Cascade Testing

As per NCCN guidelines, cascade testing of other affected and unaffected family members of the proband was performed to identify the risk of HBOC, pancreatic cancer and prostate cancer. After pre-test counselling, other family members (affected and unaffected) were tested for the previously identified *BRCA2* mutation by Sanger sequencing (i.e, capillary electrophoresis on Genetic Analyzer).

### Familial Risk Assessment

Considering the strong family history of pancreatic and breast cancer, the extended family of the proband underwent genetic testing after counselling and providing written informed consent. Among the 34 family members of the proband, six members were carriers for the identified *BRCA2* variant (c.8754G>A). These include the relatives who were previously diagnosed with hormone positive breast cancer and 3 other unaffected family members.

### Post-test Genetic Counselling

As per NCCN guidelines, the healthy unaffected *BRCA2* carriers in the extended family of the proband were counselled by genetic counsellors to explain the risk of developing HBOC. Medical advice was provided by onco-clinicians about surveillance schedules, medication, and preventive surgeries as applicable.

### Bioinformatic Analysis of *BRCA2* variant (c.8754G>A)

The *BRCA2* variant identified as c.8754G>A is located on chromosome 13q12.3. The dbSNP database has reported the identified variant as Synonymous Variant within the *BRCA2* gene (rs80359803). The ClinVar database has reported this variant (RCV000077451.3) with a clinical significance and classified as ‘pathogenic’.

This variant (c.8754G>A) resides 1 base upstream of the splice donor site within exon 21 of the *BRCA2* gene. Splicing prediction software programs such as NNSPLICE and Splice Port reveal that this variant is likely to affect splicing at the junction of exon 21 and intron 21 of the *BRCA2* gene (i.e., spliceogenic) (**Figure 2**). This spliceogenic variant was located at conserved splice donor and acceptor sites. Evolutionary conservation analysis indicates that the Exon21-Intron junction is highly conserved among multiple species. According to the ACMG pathogenic variant classification standards, spliceogenic variants are known to induce major aberrations in the transcript, generate a premature termination codon or cause in-frame deletion of a known functional domain. Therefore, the identified spliceogenic *BRCA2* variant was classified as likely pathogenic.

**Figure 1.**
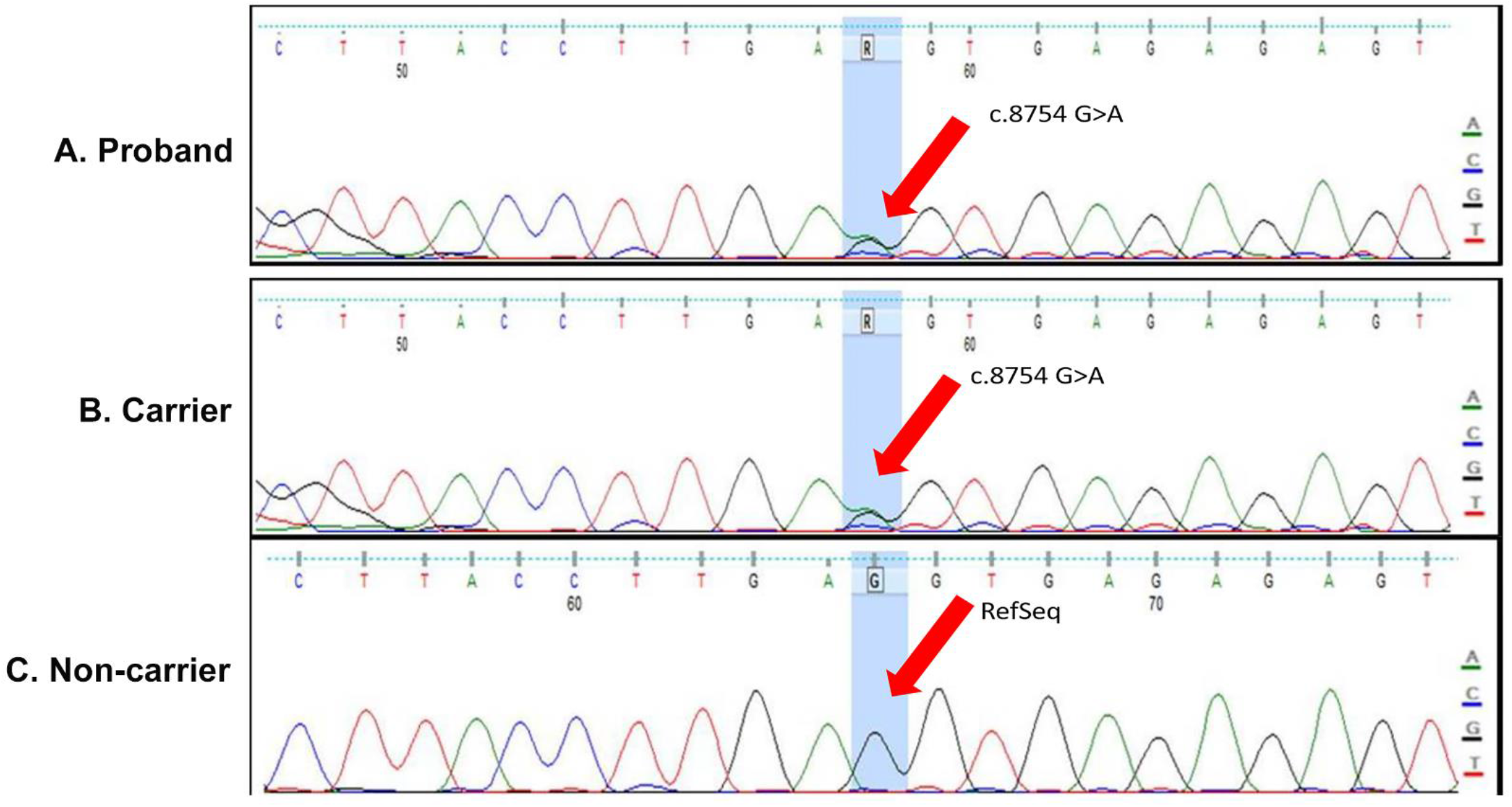
Cascade Testing. **A) Sanger sequencing data of the proband harbouring the detected variant (c**.**8754G>A)**. A heterozygous nucleotide change ‘G>A’ at position c.8754 in the *BRCA2* gene, clearly outlined on the electropherogram. **B) Sanger sequencing data revealing the individual was ‘positive’ (carrier) for the tested variant (c**.**8754G>A)**. The electropherogram clearly displays a heterozygous nucleotide change ‘G>A’ at position c.8754 in the *BRCA2* gene. **C) Sanger sequencing data revealing the individual was ‘negative’ for the tested variant (c**.**8754G>A)**. The electropherogram clearly displays no change, with reference nucleotide ‘G’ being preserved at position c.8754 in the *BRCA2* gene similar to the RefSeq.

**Figure 2.**
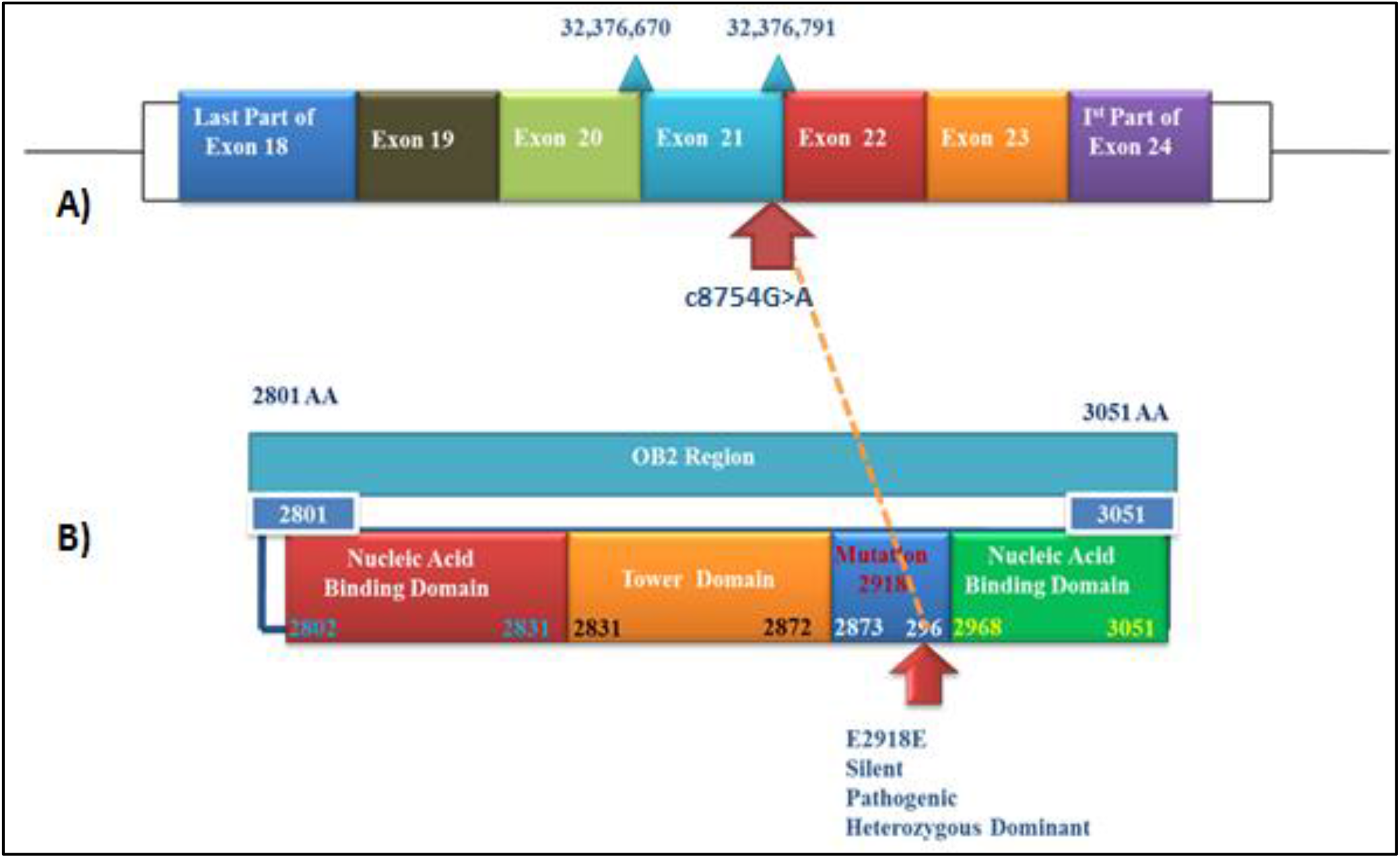
Mapping of the *BRCA2* Silent Mutation (c.8754 G>A, p.E2918E) **A)** The variant (c.8754G>A) is located 1 base upstream of the splice donor site within exon 21 of the *BRCA2* gene. **B)** Diagram representing the structure and domains of the OB2 region, contained within the DNA binding domain of the *BRCA2* gene. This region harbours the identified silent mutation (p.E2918E). Arrows indicate the position of the identified variant.

*BRCA2*, protein is a 3,418 amino acid protein constituting an N-terminal transactivation domain (23–105), 8 central BRC repeats of ∼30 amino acids each (987–2113), and a Conserved C-terminal domain *BRCA2*, CTD (2479–3152). *BRCA2* CTD has an α-helical domain and three Oligonucleotide/Oligosaccharide binding (OB) domains (OB1, OB2 and OB3). The OB2 domain (2801-3051) has multiple potential binding sites including ssDNA binding site, binding surface and interface.

At the protein level, this identified silent mutation (p.E2918E) resides in the OB2 (oligosaccharide binding fold module) region, which is contained within the DBD (DNA binding domain) of the *BRCA2* gene (**Figure 2**). The OB2 region hosting the identified BRCA2 variant is highly conserved among multiple species. *In silico* analysis did not report any change in the tertiary structure of the *BRCA2* protein. Similarly, the silent mutation was not observed to cause any change in the DNA-protein binding in wildtype and mutated form.

## DISCUSSION

In this report, we present a unique case of a male patient with pancreatic and breast cancer with a silent mutation (c.8754 G. A; p.E2918E) in the *BRCA2* gene. Over a period of 7 years starting with primary diagnosis of pancreatic cancer, the proband was diagnosed with asynchronous, bilateral, hormone positive breast cancers. Following the NCCN guidelines on genetic testing, the proband was found to be a carrier of a variant c.8754 G>A in exon 21 of the *BRCA2* gene, classified as ‘likely pathogenic’ (as per ACMG guidelines). Considering the increased risk of HBOC, prostate cancer and pancreatic cancer in family members, the affected and unaffected family members of the proband were tested for the *BRCA2* mutation by Sanger sequencing. Interestingly, two family members of the proband, diagnosed with breast cancer and three unaffected family members were also found to harbour the mutation. Even though deemed as a silent mutation (i.e., without any change in amino acids in *BRCA2* polypeptide), this mutation was clinically classified as likely pathogenic and *in silico* analysis predicted that the mutation may cause aberrant splicing of the *BRCA2* gene affecting mRNA stability.

In our report, the association of *BRCA2* pathogenic mutation (i.e., genotype) with ER positive status (i.e., phenotype) of male breast cancer is consistent with previous reports (Silvestri et al., 2016). Although not as widely studied as female breast cancers, a few studies have investigated the molecular basis of MBCs (Piscuoglio et al., 2016). Fentiman et. al. 2016 present a comprehensive review of the molecular studies on MBCs and a comparative analysis with female breast cancer cohorts. Luminal A subtype is found to be predominant in MBC with a higher *BRCA2* germline mutation frequency than breast cancer in females. Data from the CIMBA cohort analysing the characteristics of *BRCA1/2* mutations in MBCs and female breast cancers report pathologic differences (Silvestri et al., 2016). The frequency of *BRCA2* mutations in male breast cancer is found to be higher than *BRCA1* in comparison to female breast cancers (Silvestri et al., 2016). Recent studies using multigene panel testing also demonstrate differences in the molecular aetiology of MBCs (Moelans et al., 2019; Pritzlaff et al., 2017).

The most interesting finding in our study is the clinically relevant pathogenicity of the c.8754 G>A *BRCA2* variant. As a silent mutation, this variant did not induce any amino acid change in the *BRCA2* protein and yet, was classified as likely pathogenic in concordance with ACMG guidelines.

Silent mutations have been widely acknowledged to influence changes in protein expression, conformation and function (Sharma et al., 2019). Such mutations have been demonstrated to be capable of altering gene expression through mechanisms such as disruption/creation of splicing regulatory sites, gain/loss of miRNA binding sites and altering mRNA stability. Additionally, silent mutations may also affect changes in translation efficiency (translational kinetics and protein folding) by changing the codons read by tRNAs of different cellular availability (Fernández-Calero et al., 2016). Clinically relevant silent mutations have been reported within the catechol-O-methyltransferase (*COMT)* gene, an enzyme responsible for degrading catecholamines and a key regulator of pain perception and mood. The p.L136L silent mutation of *COMT* gene has shown to affect mRNA stability, subsequently having an influence on pain sensitivity and an association with mood disorders including Major depressive disorder (MDD) and bipolar disorder (BD) (Pandolfo et al., 2015). A silent mutation in the *DRD2* gene, a dopamine receptor and a key tested candidate gene in psychiatric disorders, results in a less stable secondary structure of the mRNA, further corresponding to the increased mRNA degradation and a lower protein level (Sauna & Kimchi-Sarfaty, 2011). A synonymous mutation (ΔF508) in the cystic fibrosis transmembrane conductance regulator (*CFTR)* gene has shown to alter the mRNA structure, subsequently leading to a misfolded protein (Chamary & Hurst, 2009). In cancers, such silent mutations in *ABCB1* (P-glycoprotein) gene are associated with multidrug resistance via alteration of substrate specificity (Kimchi-Sarfaty et al., 2007). Specific haplotypes of *ABCB1* (c.1236C>T, c.2677G>T and c.3435C>T) have been associated with improved treatment outcomes in patients with metastatic renal cell cancer and inflammatory bowel disease (Sauna & Kimchi-Sarfaty, 2011).

Germline *BRCA1/2* pathogenic mutations (either point mutations or gene rearrangements) are well established genetic factors associated with predisposition to HBOC. However, the contribution of silent point mutations in these key tumor suppressor genes to HBOC risk remains to be elucidated (Diederichs et al., 2016). Silent mutations in *BRCA1* gene have been demonstrated to result in reduced mRNA levels (Findlay et al., 2018). *BRCA2* silent mutations have been previously reported to result in exon skipping and changes in protein structure. The study by Hansen et. al. demonstrated that the silent mutation in exon 6 of *BRCA2* (c.744 G>A/c.516 G>A, K172K) resulted in exon 6 skipping during splicing. The resulting frameshift causes premature termination of codons 154 or 168 leading to nonsense-mediated mRNA decay, thereby potentially affecting stoichiometric amounts of protein available for biological functions.

In the current study, using a NCCN-guided clinical genetics approach for HBOC familial risk assessment, we have identified a clinically relevant *BRCA2* silent mutation (c.8754G>A, p.E2918E associated with male and female hormone positive breast cancer. In the *BRCA2* gene, this variant resides in the oligonucleotide binding (OB2) domain. This OB2 region is known to bind to DSS1, an evolutionarily conserved acidic protein, which is linked to stabilizing *BRCA2*, promoting homologous recombination and implicated in various other processes including development, and protein degradation (Pfeffer *et al*, 2017). In agreement with our bioinformatics analysis, the c.8754G>A variant leads to an alteration of the canonical donor site, and the splicing outcome leads to a 46-nt insertion of intron 21 resulting in a frameshift and premature appearance of a stop codon (Acedo et al., 2015). This identified variant was reported to induce major aberrant transcripts and thus was classified as “pathogenic” according to the class C5, five-tiered ACMG classification of variants (Acedo et al., 2015; Richards et al., 2015). This mutation and a nearby silent mutation (c.8755) has been reported in Latin American population although the prevalence in other ethnicities population is not well established (Karami & Mehdipour, 2013; Millan Catalan et al., 2019; Rebbeck et al., 2018). Also referred to as the c.8982G>A variant, this variant has been previously reported in individuals afflicted with HBOC syndrome (Finkelman et al., 2012).

Based on our familial risk profiling and *in silico* analysis of this *BRCA2* silent mutation, further functional genomics studies are warranted to understand biological mechanisms that link the genotype of *BRCA2* pathogenic variant (i.e, c.8754 G>A) to the HBOC phenotype (i.e, loss of tumor suppressor function). It is likely that such functional studies will reveal newer insights into the *BRCA2* biology with focus on mRNA stability associated with homologous recombination processes.

## Conclusion

In the current study, we report the presence of a clinically relevant silent *BRCA2* mutation (c.8754G.A, p.E2918E) characterised as likely pathogenic in six members of a family, three of whom presented with hereditary breast cancers. Although *in silico* analysis predicts formation of *BRCA2* aberrant transcripts due to this silent mutation, further mechanistic experimental studies will be needed to determine its functional role in carcinogenesis. Analysis of pathogenic silent mutations in key cancer-associated genes are expected to provide newer insights in underlying mechanisms of carcinogenesis.

## Data Availability

The study investigators would like to state that all data related to the manuscript will be available for further review on request after consultation with institutional ethics committees

## Abbreviations

BC: Breast Cancer
MBC: Male Breast Cancer
NCCN: National Comprehensive Cancer Network
HBOC: Hereditary Breast and Ovarian Cancer
USG: Ultrasonography
OB: Oligonucleotide Binding
ACMG: American College of Medical Pathologists guidelines
NGS: Next generation sequencing
gDNA: Genomic Deoxyribonucleic acid
ER: Estrogen receptor
PR: Progesterone receptor
HER2: Human epidermal growth factor receptor
CIMBA: Consortium of Investigators of Modifiers of BRCA1/2

## ACKNOWLEDGEMENT

The study authors would like to thank all participants who consented to participate in this study. We acknowledge Bajaj Auto Ltd. for providing support to research activities at Prashanti Cancer Care Mission, Pune. We are thankful to Dr Sanket Nagarkar, Dr Sneha Joshi and Nikki Dutt for their support in preparation of this manuscript.

## CONSENT

The study was approved by an Independent Ethics Committee. All subjects gave their consent for the use of their personal and medical information associated with this study.

## ETHICS APPROVAL

The study was approved by the Institutional Ethics Committee of PCCM (DCGI/CDSCO Registration Number: ECR/298/Indt/MH/2018. Every participant gave a separate informed consent for the use of their personal and medical information associated with this study. The study follows principles of the declaration of Helsinki (Brazil, 2013 version) and National Ethics Guidelines 2017 issued by the Indian Council of Medical Research, Government of India.

## References

1. Abdelwahab Yousef, A. J. (2017). Male Breast Cancer: Epidemiology and Risk Factors. Seminars in Oncology, 44(4), 267–272. https://doi.org/10.1053/j.seminoncol.2017.11.002

2. Acedo, A., Hernández-Moro, C., Curiel-García, Á., Díez-Gómez, B., & Velasco, E. A. (2015). Functional classification of BRCA2 DNA variants by splicing assays in a large minigene with 9 exons. Human Mutation, 36(2), 210–221. https://doi.org/10.1002/humu.22725

3. Chamary, J. V, & Hurst, L. D. (2009). The price of silent mutations. Scientific American, 300(6), 46–53. https://doi.org/10.1038/scientificamerican0609-46

4. Diederichs, S., Bartsch, L., Berkmann, J. C., Fröse, K., Heitmann, J., Hoppe, C., Iggena, D., Jazmati, D., Karschnia, P., Linsenmeier, M., Maulhardt, T., Möhrmann, L., Morstein, J., Paffenholz, S. V, Röpenack, P., Rückert, T., Sandig, L., Schell, M., Steinmann, A., … Wullenkord, R. (2016). The dark matter of the cancer genome: aberrations in regulatory elements, untranslated regions, splice sites, non-coding RNA and synonymous mutations. EMBO Molecular Medicine, 8(5), 442–457. https://doi.org/10.15252/emmm.201506055

5. Fentiman, I. S. (2016). Male breast cancer is not congruent with the female disease. Critical Reviews in Oncology/Hematology, 101, 119–124. https://doi.org/10.1016/j.critrevonc.2016.02.017

6. Fernández-Calero, T., Cabrera-Cabrera, F., Ehrlich, R., & Marín, M. (2016). Silent Polymorphisms: Can the tRNA Population Explain Changes in Protein Properties? Life (Basel, Switzerland), 6(1). https://doi.org/10.3390/life6010009

7. Ferzoco, R. M., & Ruddy, K. J. (2015). The Epidemiology of Male Breast Cancer. Current Oncology Reports, 18(1), 1. https://doi.org/10.1007/s11912-015-0487-4

8. Findlay, G. M., Daza, R. M., Martin, B., Zhang, M. D., Leith, A. P., Gasperini, M., Janizek, J. D., Huang, X., Starita, L. M., & Shendure, J. (2018). Accurate classification of BRCA1 variants with saturation genome editing. Nature, 562(7726), 217–222. https://doi.org/10.1038/s41586-018-0461-z

9. Finkelman, B. S., Rubinstein, W. S., Friedman, S., Friebel, T. M., Dubitsky, S., Schonberger, N. S., Shoretz, R., Singer, C. F., Blum, J. L., Tung, N., Olopade, O. I., Weitzel, J. N., Lynch, H. T., Snyder, C., Garber, J. E., Schildkraut, J., Daly, M. B., Isaacs, C., Pichert, G., … Rebbeck, T. R. (2012). Breast and ovarian cancer risk and risk reduction in Jewish BRCA1/2 mutation carriers. Journal of Clinical Oncology : Official Journal of the American Society of Clinical Oncology, 30(12), 1321–1328. https://doi.org/10.1200/JCO.2011.37.8133

10. Ibrahim, M., Yadav, S., Ogunleye, F., & Zakalik, D. (2018). Male BRCA mutation carriers: clinical characteristics and cancer spectrum. BMC Cancer, 18(1), 179. https://doi.org/10.1186/s12885-018-4098-y

11. Karami, F., & Mehdipour, P. (2013). A comprehensive focus on global spectrum of BRCA1 and BRCA2 mutations in breast cancer. BioMed Research International, 2013, 928562. https://doi.org/10.1155/2013/928562

12. Kast, K., Rhiem, K., Wappenschmidt, B., Hahnen, E., Hauke, J., Bluemcke, B., Zarghooni, V., Herold, N., Ditsch, N., Kiechle, M., Braun, M., Fischer, C., Dikow, N., Schott, S., Rahner, N., Niederacher, D., Fehm, T., Gehrig, A., Mueller-Reible, C., … Engel, C. (2016). Prevalence of BRCA1/2 germline mutations in 21 401 families with breast and ovarian cancer. Journal of Medical Genetics, 53(7), 465–471. https://doi.org/10.1136/jmedgenet-2015-103672

13. Kimchi-Sarfaty, C., Oh, J. M., Kim, I.-W., Sauna, Z. E., Calcagno, A. M., Ambudkar, S. V, & Gottesman, M. M. (2007). A ‘Silent’ Polymorphism in the *MDR* Gene Changes Substrate Specificity. Science, 315(5811), 525 LP – 528. https://doi.org/10.1126/science.1135308

14. Millan Catalan, O., Campos-Parra, A. D., Vázquez-Romo, R., Cantú de León, D., Jacobo-Herrera, N., Morales-González, F., López-Camarillo, C., Rodríguez-Dorantes, M., López-Urrutia, E., & Pérez-Plasencia, C. (2019). A Multi-Center Study of BRCA1 and BRCA2 Germline Mutations in Mexican-Mestizo Breast Cancer Families Reveals Mutations Unreported in Latin American Population. Cancers, 11(9). https://doi.org/10.3390/cancers11091246

15. Moelans, C. B., de Ligt, J., van der Groep, P., Prins, P., Besselink, N. J. M., Hoogstraat, M., Ter Hoeve, N. D., Lacle, M. M., Kornegoor, R., van der Pol, C. C., de Leng, W. W. J., Barbé, E., van der Vegt, B., Martens, J., Bult, P., Smit, V. T. H. B. M., Koudijs, M. J., Nijman, I. J., Voest, E. E., … van Diest, P. J. (2019). The molecular genetic make-up of male breast cancer. Endocrine-Related Cancer, 26(10), 779–794. https://doi.org/10.1530/ERC-19-0278

16. Nielsen, F. C., van Overeem Hansen, T., & Sørensen, C. S. (2016). Hereditary breast and ovarian cancer: new genes in confined pathways. Nature Reviews. Cancer, 16(9), 599– 612. https://doi.org/10.1038/nrc.2016.72

17. Pandolfo, G., Gugliandolo, A., Gangemi, C., Arrigo, R., Currò, M., La Ciura, G., Muscatello, M. R. A., Bruno, A., Zoccali, R., & Caccamo, D. (2015). Association of the COMT synonymous polymorphism Leu136Leu and missense variant Val158Met with mood disorders. Journal of Affective Disorders, 177, 108–113. https://doi.org/10.1016/j.jad.2015.02.016

18. Pfeffer, C. M., Ho, B. N., & Singh, A. T. K. (2017). The Evolution, Functions and Applications of the Breast Cancer Genes BRCA1 and BRCA2. Cancer Genomics & Proteomics, 14(5), 293–298. https://doi.org/10.21873/cgp.20040

19. Piscuoglio, S., Ng, C. K. Y., Murray, M. P., Guerini-Rocco, E., Martelotto, L. G., Geyer, F. C., Bidard, F.-C., Berman, S., Fusco, N., Sakr, R. A., Eberle, C. A., De Mattos-Arruda, L., Macedo, G. S., Akram, M., Baslan, T., Hicks, J. B., King, T. A., Brogi, E., Norton, L., … Reis-Filho, J. S. (2016). The Genomic Landscape of Male Breast Cancers. Clinical Cancer Research : An Official Journal of the American Association for Cancer Research, 22(16), 4045–4056. https://doi.org/10.1158/1078-0432.CCR-15-2840

20. Pritzlaff, M., Summerour, P., McFarland, R., Li, S., Reineke, P., Dolinsky, J. S., Goldgar, D. E., Shimelis, H., Couch, F. J., Chao, E. C., & LaDuca, H. (2017). Male breast cancer in a multi-gene panel testing cohort: insights and unexpected results. Breast Cancer Research and Treatment, 161(3), 575–586. https://doi.org/10.1007/s10549-016-4085-4

21. Rebbeck, T. R., Friebel, T. M., Friedman, E., Hamann, U., Huo, D., Kwong, A., Olah, E., Olopade, O. I., Solano, A. R., Teo, S.-H., Thomassen, M., Weitzel, J. N., Chan, T. L., Couch, F. J., Goldgar, D. E., Kruse, T. A., Palmero, E. I., Park, S. K., Torres, D., … Nathanson, K. L. (2018). Mutational spectrum in a worldwide study of 29,700 families with BRCA1 or BRCA2 mutations. Human Mutation, 39(5), 593–620. https://doi.org/10.1002/humu.23406

22. Richards, S., Aziz, N., Bale, S., Bick, D., Das, S., Gastier-Foster, J., Grody, W. W., Hegde, M., Lyon, E., Spector, E., Voelkerding, K., & Rehm, H. L. (2015). Standards and guidelines for the interpretation of sequence variants: a joint consensus recommendation of the American College of Medical Genetics and Genomics and the Association for Molecular Pathology. Genetics in Medicine : Official Journal of the American College of Medical Genetics, 17(5), 405–424. https://doi.org/10.1038/gim.2015.30

23. Sauna, Z. E., & Kimchi-Sarfaty, C. (2011). Understanding the contribution of synonymous mutations to human disease. Nature Reviews Genetics, 12(10), 683–691. https://doi.org/10.1038/nrg3051

24. Shah, T., Shah, N., Vijay, D. G., Patel, B., & Patel, S. (2020). Male Breast Cancer: Current Trends—a Tertiary Care Centre Experience. Indian Journal of Surgical Oncology, 11(1), 7–11. https://doi.org/10.1007/s13193-019-01021-5

25. Sharma, Y., Miladi, M., Dukare, S., Boulay, K., Caudron-Herger, M., Groß, M., Backofen, R., & Diederichs, S. (2019). A pan-cancer analysis of synonymous mutations. Nature Communications, 10(1), 2569. https://doi.org/10.1038/s41467-019-10489-2

26. Silvestri, V., Barrowdale, D., Mulligan, A. M., Neuhausen, S. L., Fox, S., Karlan, B. Y., Mitchell, G., James, P., Thull, D. L., Zorn, K. K., Carter, N. J., Nathanson, K. L., Domchek, S. M., Rebbeck, T. R., Ramus, S. J., Nussbaum, R. L., Olopade, O. I., Rantala, J., Yoon, S.-Y., … Ottini, L. (2016). Male breast cancer in BRCA1 and BRCA2 mutation carriers: pathology data from the Consortium of Investigators of Modifiers of BRCA1/2. Breast Cancer Research : BCR, 18(1), 15. https://doi.org/10.1186/s13058-016-0671-y

27. Tripoli, M., & Cordova, A. (2020). Arts and Aesthetics of Male Chest. In A. Cordova, A. Innocenti, F. Toia, & M. Tripoli (Eds.), Plastic and Cosmetic Surgery of the Male Breast (pp. 3–7). Springer International Publishing. https://doi.org/10.1007/978-3-030-25502-2_1

28. Venkitaraman, A. R. (2019). How do mutations affecting the breast cancer genes BRCA1 and BRCA2 cause cancer susceptibility? DNA Repair, 81, 102668. https://doi.org/10.1016/j.dnarep.2019.102668

29. Vietri, M. T., Caliendo, G., D’Elia, G., Resse, M., Casamassimi, A., Minucci, P. B., Cioffi, M., & Molinari, A. M. (2020). BRCA and PALB2 mutations in a cohort of male breast cancer with one bilateral case. European Journal of Medical Genetics, 63(6), 103883. https://doi.org/10.1016/j.ejmg.2020.103883

30. Weiss, J. R., Moysich, K. B., & Swede, H. (2005). Epidemiology of Male Breast Cancer. Cancer Epidemiology Biomarkers & Prevention, 14(1), 20 LP – 26. http://cebp.aacrjournals.org/content/14/1/20.abstract

